# Detection of Left Ventricular Systolic Dysfunction from Electrocardiographic Images

**DOI:** 10.1101/2022.06.04.22276000

**Authors:** Veer Sangha, Arash A Nargesi, Lovedeep S Dhingra, Akshay Khunte, Bobak J Mortazavi, Antônio H Ribeiro, Evgeniya Banina, Oluwaseun Adeola, Nadish Garg, Cynthia A Brandt, Edward J Miller, Antonio Luiz J Ribeiro, Eric J Velazquez, Luana Giatti, Sandhi M Barreto, Murilo Foppa, Neal Yuan, David Ouyang, Harlan M Krumholz, Rohan Khera

## Abstract

**Background:** Left ventricular (LV) systolic dysfunction is associated with over 8-fold increased risk of heart failure and a 2-fold risk of premature death. The use of electrocardiogram (ECG) signals in screening for LV systolic dysfunction is limited by their availability to clinicians. We developed a novel deep learning-based approach that can use ECG images for the screening of LV systolic dysfunction.

**Methods:** Using 12-lead ECGs plotted in multiple different formats, and corresponding echocardiographic data recorded within 15 days from the Yale-New Haven Hospital (YNHH) during 2015-2021, we developed a convolutional neural network algorithm to detect LV ejection fraction < 40%. The model was validated within clinical settings at YNHH as well as externally on ECG images from Cedars Sinai Medical Center in Los Angeles, CA, Lake Regional Hospital (LRH) in Osage Beach, MO, Memorial Hermann Southeast Hospital in Houston, TX, and Methodist Cardiology Clinic of San Antonia, TX. In addition, it was validated in the prospective Brazilian Longitudinal Study of Adult Health (ELSA-Brasil). Gradient-weighted class activation mapping was used to localize class-discriminating signals in ECG images.

**Results:** Overall, 385,601 ECGs with paired echocardiograms were used for model development. The model demonstrated high discrimination power across various ECG image formats and calibrations in internal validation (area under receiving operation characteristics [AUROC] 0.91, area under precision-recall curve [AUPRC] 0.55), and external sets of ECG images from Cedars Sinai (AUROC 90, AUPRC 0.53), outpatient YNHH clinics (AUROC 0.94, AUPRC 0.77), LRH (AUROC 0.90, AUPRC 0.88), Memorial Hermann Southeast Hospital (AUROC 0.91, AUPRC 0.88), Methodist Cardiology Clinic (AUROC 0.90, AUPRC 0.74), and ELSA-Brasil cohort (AUROC 0.95, AUPRC 0.45). An ECG suggestive of LV systolic dysfunction portended over 27-fold higher odds of LV systolic dysfunction on TTE (OR 27.5, 95% CI, 22.3-33.9 in the held-out set). Class-discriminative patterns localized to the anterior and anteroseptal leads (V2-V3), corresponding to the left ventricle regardless of the ECG layout. A positive ECG screen in individuals with LV ejection fraction _≥_ 40% at the time of initial assessment was associated with a 3.9-fold increased risk of developing incident LV systolic dysfunction in the future (HR 3.9, 95% CI 3.3-4.7, median follow-up 3.2 years).

**Conclusions:** We developed and externally validated a deep learning model that identifies LV systolic dysfunction from ECG images. This approach represents an automated and accessible screening strategy for LV systolic dysfunction, particularly in low-resource settings.

**CLINICAL PERSPECTIVE:** *What is New?:*

- A convolutional neural network model that accurately identifies LV systolic dysfunction from ECG images across subgroups of age, sex, and race.
- The model shows robust performance across multiple institutions and health settings, both applied to ECG image databases as well as directly uploaded single ECG images to a web-based application by clinicians.
- The approach provides information for both screening of LV systolic dysfunction and its risk based on ECG images alone.

*What are the clinical implications?:*

- Our model represents an automated screening strategy for LV systolic dysfunction on a variety of ECG layouts.
- With availability of ECG images in practice, this approach overcomes implementation challenges of deploying an interoperable screening tool for LV systolic dysfunction in resource-limited settings.
- This model is available in an online format to facilitate real-time screening for LV systolic dysfunction by clinicians.

## INTRODUCTION

Left ventricular (LV) systolic dysfunction is associated with over 8-fold increased risk of subsequent heart failure and nearly 2-fold risk of premature death.^1^ While early diagnosis can effectively lower this risk,^2–4^ individuals are often diagnosed after developing symptomatic disease due to lack of effective screening strategies.^5–7^ The diagnosis traditionally relies on echocardiography, a specialized imaging modality that is resource intensive to deploy at scale.^8,9^ Algorithms using raw signals from electrocardiography (ECG) have been developed as a strategy to detect LV systolic dysfunction.^10–12^ However, clinicians, particularly in remote settings, do not have access to ECG signals. The lack of interoperability in signal storage formats from ECG devices further limits the broad uptake of such signal-based models.^13^ The use of ECG images is an opportunity to implement interoperable screening strategies for LV systolic dysfunction.

We previously developed a deep learning approach of format-independent inference from real-world ECG images.^14^ The approach can interpretably diagnose cardiac conduction and rhythm disorders using any layout of real-world 12-lead ECG images and can be accessed on web- or application-based platforms. Extension of this artificial intelligence (AI)-driven approach to ECG images to screen for LV systolic dysfunction could rapidly broaden access to a low cost, easily accessible, and scalable diagnostic approach to underdiagnosed and undertreated at-risk populations. This approach adapts deep learning for end-users, without disruption of data pipelines or clinical workflow. Moreover, the ability to add localization of predictive cues in the ECG images relevant to the LV can improve the uptake of these models in clinical practice.^15^

In this study, we present a model for accurate identification of LV ejection fraction (LVEF) less than 40%, a threshold with therapeutic implications, based on ECG images. We developed, tested, and externally validated this approach using paired ECG-echocardiographic data from large academic hospitals, rural hospital systems, and a prospective cohort study.

## METHODS

The Yale Institutional Review Board reviewed the study, which approved the study protocol and waived the need for informed consent as the study represents a secondary analysis of existing data. The data cannot be shared publicly though an online version of the model is publicly available for research use at https://www.cards-lab.org/ecgvision-lv.

### Data Source for Model Development

We used 12-lead ECG signal waveform data from the Yale New Haven Hospital (YNHH) collected between 2015 and 2021. These ECGs were recorded as standard 12-lead recordings sampled at a frequency of 500 Hz for 10 seconds. These were recorded on multiple different machines and a majority were collected using Philips PageWriter machines and GE MAC machines. Among patients with an ECG, those with a corresponding transthoracic echocardiogram (TTE) within 15 days of obtaining the ECG were identified from the YNHH electronic health records. LVEF values were extracted based on a cardiologist’s read of the nearest TTE to each ECG. To augment the evaluation of models built on an image dataset generated from this YNHH signal waveform, six sets of ECG image datasets were used for external validation.

### Data Preprocessing

All ECGs were analyzed to determine whether they had 10 seconds of continuous recordings across all 12 leads. The 10 second samples were preprocessed with a one second median filter, which was subtracted from the original waveform to remove baseline drift in each lead, representing processing steps pursued by ECG machines before generating printed output from collected waveform data.

ECG signals were transformed into ECG images using the python library ecg-plot,^16^ and stored at 100 DPI. Images were generated with a calibration of 10 mm/mV, which is standard for printed ECGs in most real-world settings. In sensitivity analyses, we evaluated model performance on images calibrated at 5 and 20 mm/mV. All images, including those in train, validation, and test sets, were converted to greyscale, followed by down-sampling to 300×300 pixels regardless of their original resolution using Python Image Library (PIL v9.2.0). To ensure that the model was adaptable to real-world images, which may vary in formats and the layout of leads, we created a dataset with different plotting schemes for each signal waveform recording (**Figure 1**). This strategy has been used to train a format-independent image-based model for detecting conduction and rhythm disorders as well as the hidden label of gender.^14^ The model in this study learned ECG lead-specific information based on the label regardless of the location of the lead.

**Figure 1.**
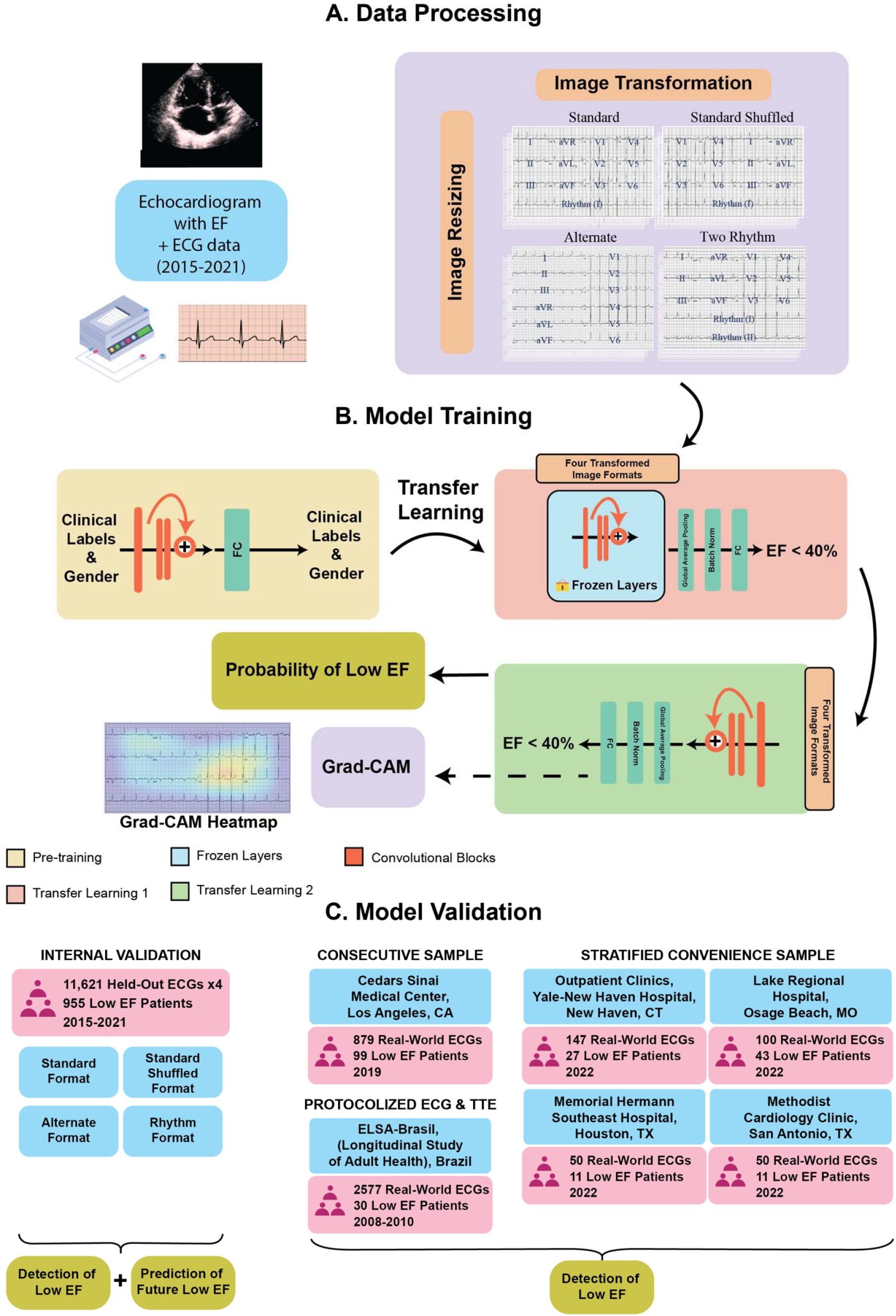
Study Outline A) Data processing, B) Model training, and C) Model validation. Abbreviations: ECG, electrocardiogram; EF, ejection fraction; FC, fully connected layers; Grad-CAM, gradient-weighted class activation mapping; CT, Connecticut; ELSA-Brasil, Estudo Longitudinal de Saúde do Adulto (The Brazilian Longitudinal Study of Adult Health); MO, Missouri; TX, Texas. *We pursued a transfer learning strategy in developing the current model from our previous algorithm which was originally trained to detect cardiac rhythm disorders and the hidden label of gender from ECG images. The transfer learning was used as initialization weights for the EfficientNet B3 convolutional neural network being trained to detect LV systolic dysfunction. Other than the weights, clinical and gender labels were not input to the current model.

Four formats of images were included in the training image dataset (**Figure 1**). The first format was based on the standard printed ECG format in the United States, with four 2.5 second columns printed sequentially on the page. Each column contained 2.5 second intervals from three leads. The full 10-second recording of the lead I signal was included as the rhythm strip. The second format, a two-rhythm format, added lead II as an additional rhythm strip to the standard format. The third layout was the alternate format which consisted of two columns, the first with six simultaneous 5-second recordings from the limb leads, and the second with six simultaneous 5-second recordings from the precordial leads, without a corresponding rhythm lead. The fourth format was a shuffled format, which had precordial leads in the first two columns and limb leads in the third and fourth. All images were rotated a random amount between -10 and 10 degrees before being input into the model to mimic variations seen in uploaded ECGs and to aid in prevention of overfitting.

The process of converting ECG signals to images was independent of model development, ensuring that the model did not learn any aspects of the processing that generated images from the signals. All ECGs were converted to images in all different formats without conditioning on clinical labels. The validation required uploaded images to be upright, cropped to the waveform region, with no brightness and contrast consideration as long as the waveform is distinguishable from the background and lead labels are discernible.

### Experimental Design

Each included ECG had a corresponding LVEF value from its nearest TTE within 15 days of recording. Low LVEF was defined as LVEF < 40%, the cutoff used as an indication for most guideline-directed pharmacotherapy for heart failure.^4^ Patients with at least one ECG within 15 days of its nearest TTE were randomly split into training, validation, and held-out test patient level sets (85%, 5%, 10%, **Figure S1**). This sampling was stratified by whether a patient had ever had LVEF < 40% to ensure cases of preserved and reduced LVEF were split proportionally among the sets. In the training cohort, all ECGs within 15 days of a TTE were included for all patients to maximize the data available. In validation and testing cohorts, only one ECG was included per patient to ensure independence of observations in the assessment of performance metrics. This ECG was randomly chosen amongst all ECGs within 15 days of a TTE. Additionally, to ensure that model learning was not affected by the relatively lower frequency of LVEF < 40%, higher weights were given to these cases at the training stage based on the effective number of samples class sampling scheme.^17^

### Model Training

We built a convolutional neural network model based on the EfficientNet-B3 architecture,^18^ which previously demonstrated an ability to learn and identify both rhythm and conduction disorders, as well as the hidden label of gender in real-world ECG images.^14^ The EfficientNet-B3 model requires images to be sampled at 300 × 300 square pixels, includes 384 layers, and has over 10 million trainable parameters (**Figure S2**). We utilized transfer learning by initializing model weights as the pretrained EfficientNet-B3 weights used to predict the six physician-defined clinical labels and gender from Sangha et al.^14^ We first only unfroze the last four layers and trained the model with a learning rate of 0.01 for 2 epochs, and then unfroze all layers and trained with a learning rate of 5 × 10^−6^ for 6 epochs. We used an Adam optimizer, gradient clipping, and a minibatch size of 64 throughout training. The optimizer and learning rates were chosen after hyperparameter optimization. For both stages of training the model, we stopped training when validation loss did not improve in 3 consecutive epochs.

We trained and validated our model on a generated image dataset that had equal numbers of standard, two-rhythm, alternate, and standard shuffled images (**Figure 1**). In sensitivity analyses, the model was validated on three novel ECG layouts constructed from the held-out set to assess its performance on ECG formats not encountered in the training process. These novel ECG outlines included three-rhythm (with leads I, II, and V1 as the rhythm strip), no rhythm, and rhythm on top formats (with lead I as the rhythm strip located above the 12-lead, **Figure S3**). Additional sensitivity analyses were performed using ECG images calibrated at 5, 10, and 20 mm/mV (**Figure S4**). A custom class-balanced loss function (weighted binary cross-entropy) based on the effective number of samples was used given the lower frequency of the LVEF < 40% label relative to those with an LVEF _≥_ 40%.

### External validation

We pursued a series of validation studies. These represented both clinical and population-based cohort studies. Clinical validation represented non-synthetic image datasets from clinical settings spanning (1) consecutive patients undergoing outpatient echocardiography at the Cedars Sinai Medical Center in Los Angeles, CA, and (2) stratified convenience samples of LV systolic dysfunction and non-LV systolic dysfunction ECGs from four different settings (a) outpatient clinics of YNHH, (b) inpatient admissions at Lake Regional Hospital (LRH) in Osage Beach, MO, (c) inpatient admissions at Memorial Hermann Southeast Hospital in Houston, TX, (d) outpatient visits and inpatient admissions at Methodist Cardiology Clinic in San Antonio, TX. In addition, we validated our approach in the prospective cohort from Brazil, the Brazilian Longitudinal Study of Adult Health (ELSA-Brasil),^19^ with protocolized ECG and echocardiogram in study participants.

Inclusion and exclusion criteria for external validation sets were similar to the internal YNHH dataset. Patients were limited to those having a 12-lead ECG within 15 days of a TTE with reported LVEF. For patients with more than one TTE in this interval, the LVEF from the nearest TTE was used for analysis.

At Cedars Sinai, all index ECG images from consecutive patients undergoing outpatient visits between, January through March 2019, representing 879 individuals, including 99 with LVEF < 40%, were included. These analyses were performed in a fully federated and blinded fashion without access to any of the ECG data to the algorithm’s developers.

For the other clinical validation sites, a stratified convenience sample enriched for low LVEF was drawn. This was done to evaluate the broad use in a clinical setting by practicing clinicians without access to a research dataset. Our preliminary assessment of LV systolic dysfunction prevalence in outpatient and inpatient settings were 10% and 20%, respectively. We sought to achieve twice this prevalence in our external validation data in these sites to ensure our performance was not driven by patients with preserved LVEF and that the model could detect those with LV systolic dysfunction. Specifically, a 1:4 ratio of ECGs corresponding to LVEF < 40% and _≥_ 40% was sought at three of the four sites (YNHH, Memorial Hermann Southeast Hospital, and Methodist Cardiology Clinic). At the fourth site, LRH, a 1:2 ratio was requested to better measure the model’s discriminative ability in an inpatient-only setting.

In addition to the clinical validation studies, where concurrent ECG and echocardiogram are always clinically indicated, imposing a selection of the population, we evaluated our model in the ELSA-Brasil study, a community-based prospective cohort in Brazil that obtained ECG and echocardiography from participants on the enrollment visit between 2008-2010. This set included data from 2,577 individuals, including 30 from individuals with LVEF < 40%.

Before validation, patient identifiers, ECG measurements, and reported diagnoses were removed from all ECG images. The differences in ECG layouts and the procedures for validation are described in further detail in the **Online Supplement**. Deidentified samples of ECG images are presented in **Figure S5** (Cedars Sinai Medical Center), **Figure S6** (YNHH and LRH), **Figure S7** (Memorial Hermann Southeast Hospital), and **Figure S8** (Methodist Cardiology Clinic), and images are available from the authors upon request.

### Localization of Model Predictive Cues

We used Gradient-weighted Class Activation Mapping (Grad-CAM) to highlight which portions of an image were important for the prediction of LVEF < 40%.^20^ We calculated the gradients on the final stack of filters in our EfficientNet-B3 model for each prediction and performed a global average pooling of the gradients in each filter, emphasizing those that contributed to a prediction.

We then multiplied these filters by their importance weights and combined them across filters to generate Grad-CAM heatmaps, which we overlayed on the original ECG images. We averaged class activation maps among 100 positive cases with the most confident model predictions for LVEF < 40% across ECG formats to determine the most important image areas for the prediction of low LVEF. We took an arithmetic mean across the heatmaps for a given image format, and overlayed this average heatmap across a representative ECG to understand it in context. The activation map, a 10×10 array was upsampled to the original image size using the bilinear interpolation built into TensorFlow v 2.8.0. We also evaluated the Grad-CAM for individual ECGs to evaluate the consistency of the information on individual examples.

### Statistical Analysis

Categorical variables were presented as frequency and percentages, and continuous variables as means and standard deviations or median and interquartile range, as appropriate. Model performance was evaluated in the held-out test set and external ECG image datasets. We used area under the receiver operator characteristic (AUROC) to measure model discrimination. The cut-off for binary prediction of LV systolic dysfunction was set at 0.10 for all internal and external validations, based on the threshold that achieved a sensitivity of over 90% in the internal validation set. We also assessed area under precision recall curve (AUPRC), sensitivity, specificity, positive predictive value (PPV), negative predictive value (NPV), and diagnostic odds ratio. 95% CIs for AUROC and AUPRC were calculated using DeLong’s algorithm and bootstrapping with 1000 variations for each estimate, respectively.^21,22^ Model performance was assessed across demographic subgroups and ECG outlines, as described above. We conducted further sensitivity analyses of model performance across ECG calibrations, PR intervals, and after excluding paced rhythms, conduction disorders, atrial fibrillation, and atrial flutter. Moreover, we assessed the association of the model’s predicted probability of LV systolic dysfunction across LVEF categories.

Next, we evaluated the future development of LV systolic dysfunction in time-to-event models using a Cox proportional hazards model. In this analysis, we took the first temporal ECG from the patients in the held-out test set, and then modeled the first development of LVEF < 40% across the groups of patients who screened positive but did not have concurrent LV systolic dysfunction (false positives), and those that screened negative (true negative) from this first ECG, with censored at death or end of study period in June 2021. Additionally, we computed an adjusted hazard ratio that accounted for differences in age, sex, and baseline LVEF at the time of index screening for visualization of survival trends. Analytic packages used in model development and statistical analysis are reported in **Table S1**. All model development and statistical analyses were performed using Python 3.9.5 and the level of significance was set at an alpha of 0.05.

## RESULTS

### Study Population

Out of the 2,135,846 ECGs obtained between 2015 to 2021, 440,072 were from patients who had TTEs within 15 days of obtaining the ECG. Overall, 433,027 had a complete ECG recording, representing 10 seconds of continuous recordings across all 12 leads. These ECGs were drawn from 116,210 unique patients and were split into train, validation, and test sets at a patient level (**Figure S1**).

A total of 116,210 individuals with 385,601 ECGs constituted the study population, representing those included in the train, validation, test sets. Individuals whose ECGs were used for model development had a median age of 68 years (IQR 56, 78) at the time of ECG recording, and 59,282 (51.0%) were women. Overall, 75,928 (65.3%) were non-Hispanic white, 14,000 (12.0%) non-Hispanic Black, 9,349 (8.0%) Hispanic, and 16,843 (14.5%) were from other races. A total of 56,895 (14.8%) ECGs had a corresponding echocardiogram with an LVEF below 40%, 36,669 (9.5%) had an LVEF greater than or equal to 40% but less than 50%, and 292,037 (75.7%) had LVEF 50% or greater (**Table S2)**.

### Detection of LV Systolic Dysfunction

The model’s AUROC for detecting LVEF < 40% on the held-out test set composed of standard images was 0.91 and its AUPRC was 0.55 **(Figure 2**). A probability threshold for predicting LVEF < 40% was chosen based on a sensitivity of 0.90 or higher in the validation subset. With this threshold, the model had sensitivity and specificity of 0.89 and 0.77 in the held-out test set, and PPV and NPV of 0.26 and 0.99, respectively. Overall, an ECG suggestive of LV systolic dysfunction portended over 27-fold higher odds (OR 27.5, 95% CI, 22.3 – 33.9) of LV systolic dysfunction on TTE (**Table 1**). The model’s performance was comparable across subgroups of age, sex, and race (**Table 1** and **Figure 2**). Moreover, across successive deciles of the model predicted probabilities, the proportion of individuals with LV systolic dysfunction increased, while the mean LVEF decreased **(Figure S9)**.

**Figure 2.**
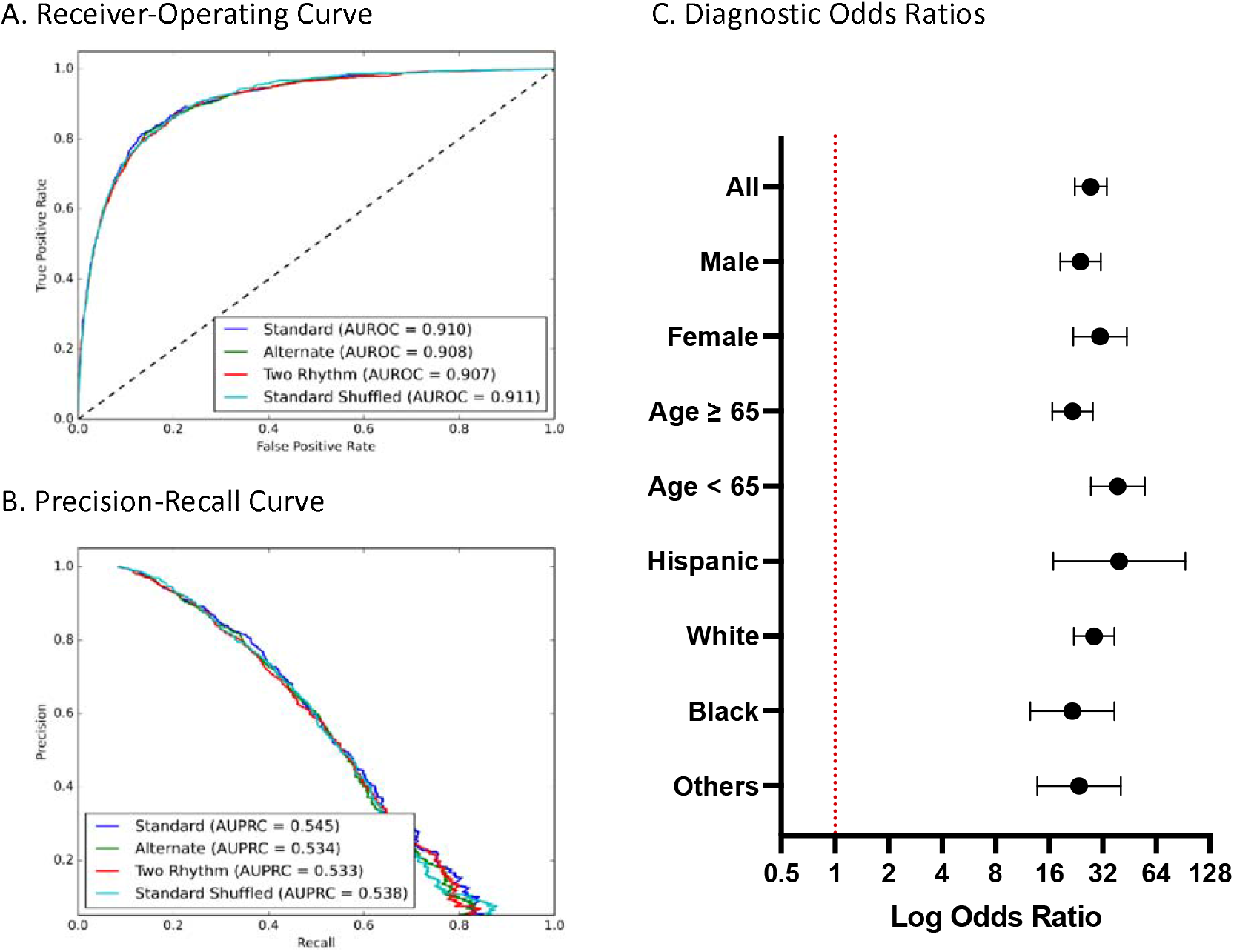
Model Performance Measures A) Receiver-Operating and B) Precision-Recall Curves on images in held-out test set C) Diagnostic Odds Ratios across age, gender, and race subgroups on standard format images in the held-out test set. Abbreviations: AUROC, area under receiver-operating characteristic curve; AUPRC, area under precision-recall curve.

**Table 1.** Performance of model on test images across demographic subgroups in the held-out test set. Abbreviations: PPV, positive predictive value; NPV, negative predictive value; AUROC, area under receiver operating characteristic curve; AUPRC, area under precision recall curve; OR, odds ratio

| Labels | Number | PPV | NPV | Specificity | Sensitivity | AUROC | AUPRC |
| --- | --- | --- | --- | --- | --- | --- | --- |
| **All** | 11621<br>(100%) | 0.257 | 0.988 | 0.769 | 0.892 | 0.910 (0.901-0.919) | 0.545 (0.511 – 0.579) |
| **Male** | 5952<br>(51.2%) | 0.285 | 0.984 | 0.735 | 0.897 | 0.901 (0.889-0.914) | 0.583 (0.539 – 0.621) |
| **Female** | 5668<br>(48.8%) | 0.215 | 0.991 | 0.802 | 0.884 | 0.917 (0.903-0.932) | 0.470 (0.416 – 0.530) |
| **≥ 65** | 6550<br>(56.4%) | 0.252 | 0.985 | 0.717 | 0.896 | 0.892 (0.880-0.905) | 0.522 (0.480 – 0.561) |
| **< 65** | 5068<br>(43.6%) | 0.266 | 0.991 | 0.833 | 0.886 | 0.931 (0.916-0.945) | 0.590 (0.534 – 0.655) |
| **Hispanic** | 942<br>(8.1%) | 0.253 | 0.992 | 0.802 | 0.908 | 0.926 (0.892-0.961) | 0.576 (0.453 – 0.696) |
| **White** | 7557<br>(65.0%) | 0.261 | 0.988 | 0.770 | 0.895 | 0.910 (0.898-0.921) | 0.537 (0.498 – 0.580) |
| **Black** | 1417<br>(12.2%) | 0.263 | 0.984 | 0.712 | 0.897 | 0.899 (0.872-0.925) | 0.590 (0.498 – 0.665) |
| **Other** | 1705<br>(14.7%) | 0.231 | 0.987 | 0.787 | 0.864 | 0.912 (0.887-0.937) | 0.532 (0.437 – 0.625) |

\* Gender information was not available for 1 patient and age was not available for 3 patient of the total 11,621 patients in the held-out test set

### Model Performance Across ECG Formats and Calibrations

The model performance was comparable across the four original layouts of ECG images in the held-out set with AUROC of 0.91 in detecting concurrent LV systolic dysfunction (**Table S3**). The model had a sensitivity of 0.89 and a positive prediction conferred 26- to 27-fold higher odds of LV systolic dysfunction on the standard and the three variations of the data. In sensitivity analyses, the model demonstrated similar performance in detecting LV systolic dysfunction from novel ECG formats that were not encountered before, with AUROC between 0.88-0.91 **(Table S4)**.

The model performance was also consistent across ECG calibrations with an AUROC between 0.88 and 0.91 on ECG calibrations of 5, 10, and 20 mm/mV and AUROC 0.908 (0.899 – 0.918) and AUPRC of 0.538 (0.503 – 0.573) with mixed calibrations in the held-out test set. The mixed calibration was generated with a random sample of 5 mm/mV and 20 mm/mV calibrations from the highest and lowest quartiles of voltages, respectively, in lead I (together representing 25% of the sample from the test set), along with 10 mm/mV (remaining 75% of test set) (**Table S5**). Further sensitivity analyses demonstrated consistent model performance on ECGs (a) without prolonged PR interval (AUROC 0.920 and AUPRC 0.537, **Table S6**), (b) without paced rhythms (AUROC 0.908, AUPRC 0.519, **Table S7**), and (c) without atrial fibrillation, atrial flutter, and conduction disorders (AUROC 0.919, AUPRC 0.536, **Table S8**). Model performance was also consistent across subsets on the held-out test set based on the timing of the ECG relative to the echocardiogram (**Table S9**).

### LV Systolic Dysfunction in Model-predicted False Positives

Of the 10,666 ECGs in the held-out test set with an associated LVEF _≥_ 40% on a proximate echocardiogram, the model classified 2,469 (23.1%) as “false positives”, and 8,197 (76.9%) as true negatives. In further evaluation of false positives, 562 (22.8% of false positives) had evidence of mild LV systolic dysfunction with LVEF between 40-50% on concurrent echocardiography.

In this group of individuals, 4,046 patients had at least one follow-up TTE, including 1,125 (27.8%) false positives and 2,921 (72.2%) true negatives on the initial index screen. There were 2,665 and 6,083 echocardiograms in the false positive and true negative populations during the follow-up, with the longest follow-up of 6.1 years. Overall, 264 (23.5%) patients with model-predicted positive screen and 199 (6.8%) with negative screen developed new LVEF < 40% over the median follow-up of 3.2 years (IQR 1.8-4.4 years, **Figure 3**). This represented a 3.9-fold higher risk of incident low LVEF based on having a positive screening result (HR 3.9, 95% CI 3.3-4.7). After adjustment for age, sex, and LVEF at the time of screening, patients with positive screen had a 2.3-fold higher risk of incident low LVEF (Adjusted HR 2.3, 95% CI 1.9-2.8).

**Figure 3.**
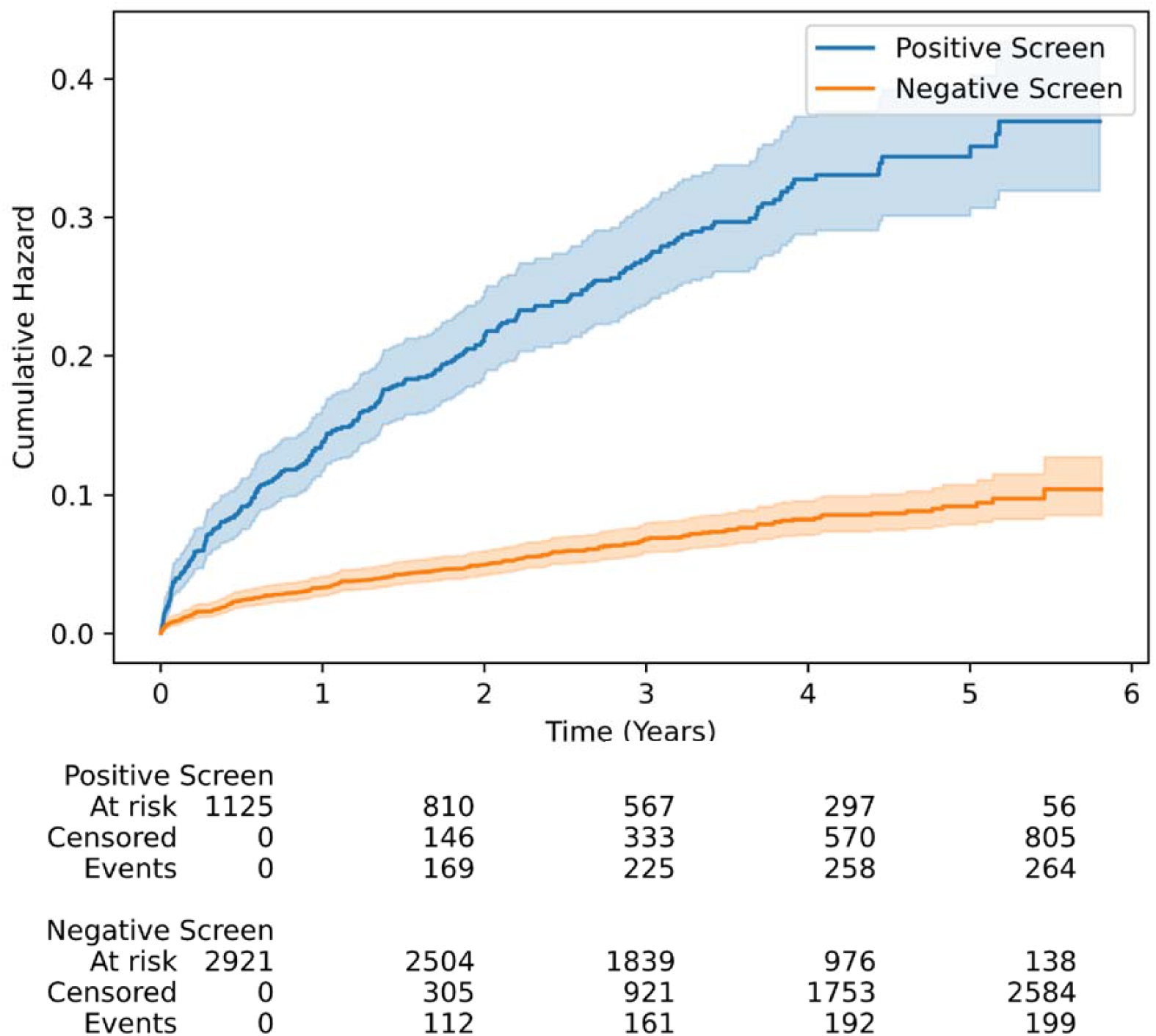
Cumulative hazard curves for incident LV systolic dysfunction in model-predicted positive and negative screens amongst the members of the held-out test set with LVEF ≥ 40% and at least one follow-up measurement.

### Localization of Predictive Cues for LV Systolic Dysfunction

Class activation heatmaps of the 100 positive cases with the most confident model predictions for reduced LVEF prediction across four ECG layouts are presented in **Figure 4**. For all four formats of images, the region corresponding to leads V2 and V3 were the most important areas for prediction of reduced LVEF. Representative images of Grad-CAM analysis in sampled individuals with positive and negative screens in the held-out test set, and non-synthetic ECG images in validation sites are presented in **Figures S10** and **S11**, respectively.

**Figure 4.**
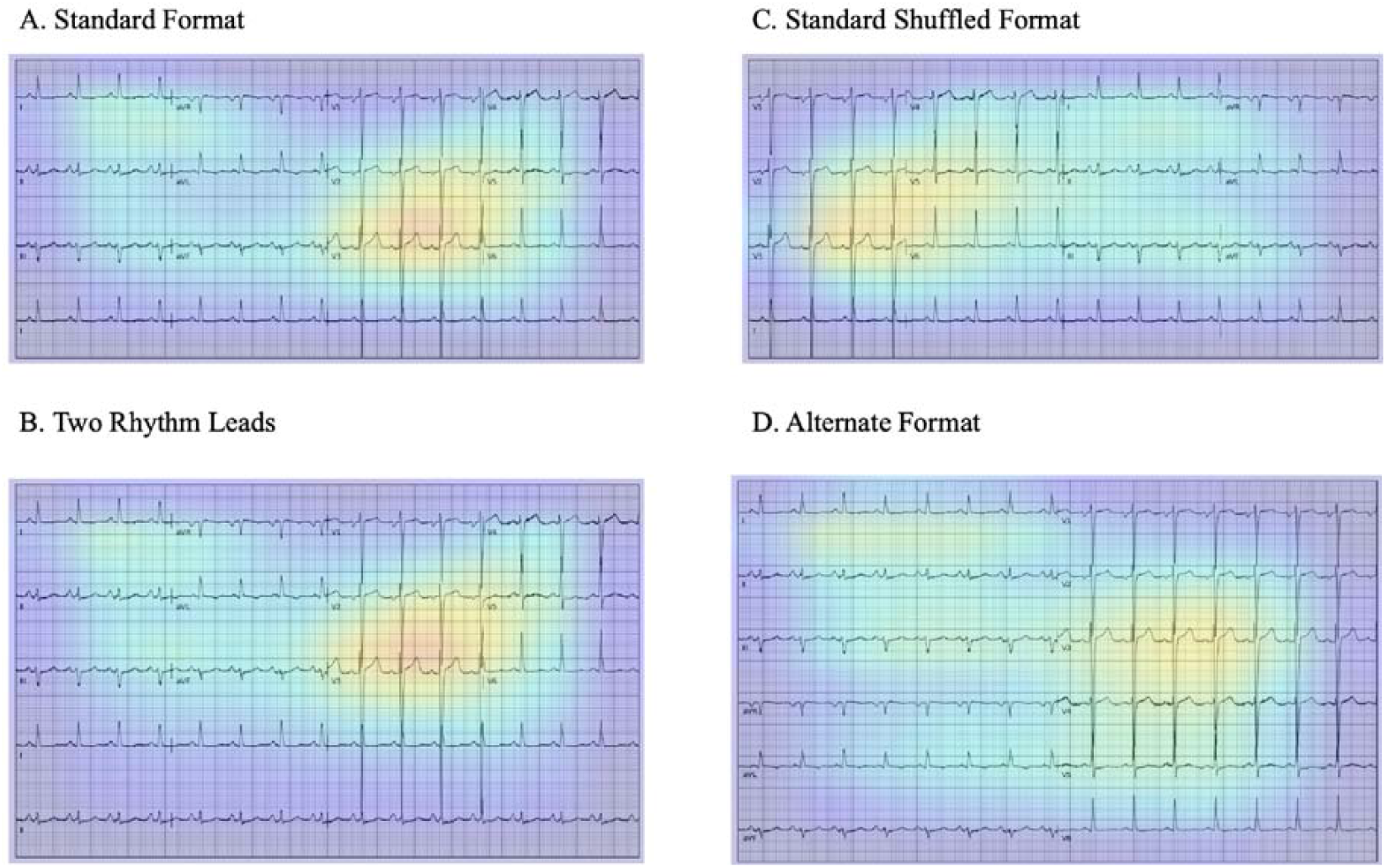
Gradient-weighted Class Activation Mapping (Grad-CAMs) across ECG formats. A) Standard format B) Two rhythm leads C) Standard shuffled format D) Alternate format. The heatmaps represent averages of the 100 positive cases with the most confident model predictions for LVEF < 40%.

### External Validation

The validation performance of the model was consistent and robust across each of the 6 validation datasets (**Figure 5**). The first validation set at Cedars Sinai Medical Center included 879 ECGs from consecutive patients who underwent outpatient echocardiography, including 99 (11%) individuals with LVEF < 40%. The model demonstrated an AUROC of 0.90 and an AUPRC of 0.53 in this set. Second, a total of 147 ECG images drawn from YNHH outpatient clinics were used for validation and included 27 images (18%) from patients with LVEF < 40%. The model had an AUROC of 0.94 and AUPRC of 0.77 in validation on these images. The third image dataset included ECG images from inpatient visits to the LRH. It included 100 ECG images, with 43 images (43%) from patients with LVEF < 40%, with a model AUROC of 0.90 and AUPRC of 0.88. The fourth dataset from Memorial Hermann Southeast Hospital included 50 ECG images, 11 (22%) from patients with LVEF < 40%, with a model AUROC and AUPRC of 0.91 and 0.88 on these images, respectively. The fifth validation set contained 50 ECG images from the Methodist Cardiology Clinic, which included 11 (20%) ECGs from patients with LVEF < 40%, with model AUROC of 0.90 and AUPRC of 0.74.

**Figure 5.**
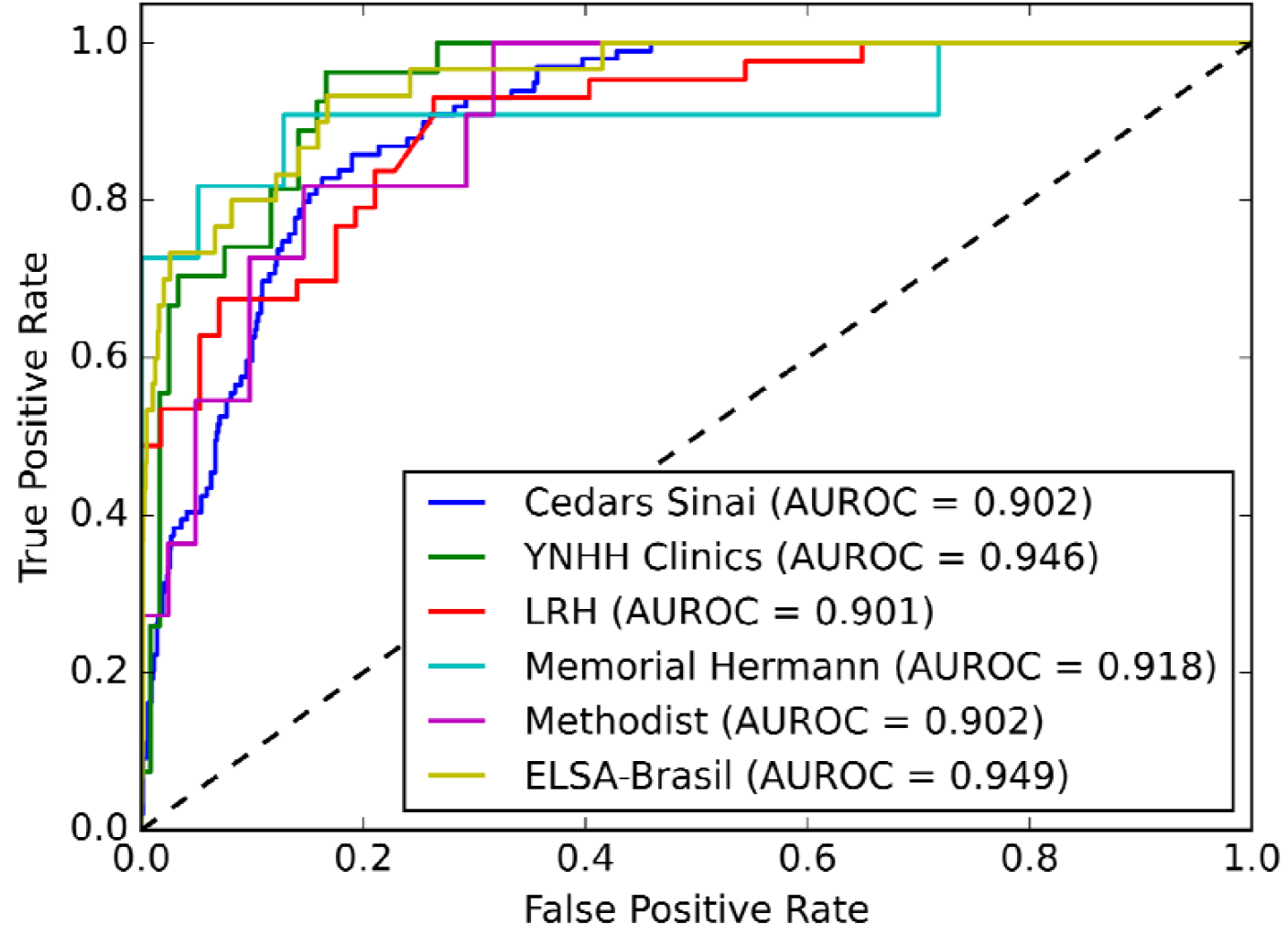
Receiver-Operating Curves for external validation sites. Abbreviations: AUROC, area under receiver-operating characteristic curve; EF, Ejection fraction; LRH, Lake Regional Hospital; YNHH, Yale New Haven Hospital

The sixth set included 2,577 ECGs from prospectively enrolled individuals in the ELSA-Brasil study, including 30 with LVEF < 40%. The model demonstrated an AUROC 0.95 and AUPRC 0.45 on this set. The model performance on these 6 validation sets is outlined in **Table 2** and **Table S10**.

**Table 2.** Performance of model on external validation datasets. Abbreviations: PPV, positive predictive value; NPV, negative predictive value; AUROC, area under receiver operating characteristic curve; AUPRC, area under precision recall curve; ELSA-Brasil, Estudo Longitudinal de Saúde do Adulto (The Brazilian Longitudinal Study of Adult Health)

| Site | PPV | NPV | Specificity | Sensitivity | AUROC | AUPRC |
| --- | --- | --- | --- | --- | --- | --- |
| Cedars Sinai Medical Center | 0.326 | 0.979 | 0.772 | 0.869 | 0.902 (0.877 – 0.926) | 0.533 (0.432 – 0.640) |
| Outpatient Clinics of YNHH | 0.338 | 1.000 | 0.558 | 1.000 | 0.946 (0.910 - 0.982) | 0.775 (0.605 – 0.916) |
| LRH | 0.538 | 0.955 | 0.368 | 0.977 | 0.901 (0.843 - 0.959) | 0.889 (0.810 – 0.946) |
| Memorial Hermann Southeast Hospital | 0.385 | 0.958 | 0.590 | 0.909 | 0.918 (0.790 - 1.000) | 0.888 (0.699 – 1.000) |
| Methodist Cardiology Clinic | 0.458 | 1.000 | 0.667 | 1.000 | 0.902 (0.816 - 0.989) | 0.738 (0.470 – 0.928) |
| ELSA-Brasil | 0.256 | 0.996 | 0.976 | 0.700 | 0.949 (0.915 – 0.983) | 0.449 (0.290 – 0.651) |

## DISCUSSION

We developed and externally validated an automated deep learning algorithm that accurately identifies LV systolic dysfunction solely from ECG images. The algorithm has high discrimination and sensitivity, representing characteristics ideal for a screening strategy. It is robust to variations in the layouts of ECG waveforms and detects the location of ECG leads across multiple formats with consistent accuracy, making it suitable for implementation in a variety of settings. Moreover, the algorithm was developed and tested in a diverse population with high performance in subgroups of age, sex, and race, and across geographically dispersed academic and community health systems. It performed well in 6 external validation sites, spanning both clinical settings as well as a prospective cohort study where protocolized echocardiograms were performed concurrently with ECGs. An evaluation of the class-discriminating signals localized it to the anteroseptal and anterior leads regardless of the ECG layout, topologically corresponding to the left ventricle. Finally, among individuals who did not have a concurrently recorded low LVEF, a positive ECG screen was associated with a 3.9-fold increased risk of developing LV systolic dysfunction in the future compared with those with negative screen, which was significant after adjustment for age, sex, and baseline LVEF. Therefore, an ECG image-based approach can represent a screening as well as predictive strategy for LV systolic dysfunction, particularly in low-resource settings.

Image-based analysis of ECGs through deep learning represents a novel application of AI to improve clinical care. Convolutional neural networks have previously been designed to detect low LVEF from ECG signals.^10,11^ Although reliance of signal-based models on voltage data is not computationally limited, their use in both retrospective and prospective settings requires access to a signal repository where the ECG data architecture varies by ECG device vendors.

Moreover, data are often not stored beyond generating printed ECG images, particularly in remote settings.^23^ Furthermore, widespread adoption of signal-based models is limited by the implementation barriers requiring health system-wide investments to incorporate them into clinical workflow, something that may not be available or cost-effective in low-resource settings and, to date, is not widely available in higher resource setting such as the US. The algorithm reported in this study overcomes these limitations by making detection of LV systolic dysfunction from ECGs interoperable across acquisition formats and directly available to clinicians who only have access to ECG images. Since scanned ECG images are the most common format of storage and use of electrocardiograms, untrained operators can implement large scale screening through chart review or automated applications to image repositories – a lower resource task than optimizing tools for different machines.

The use of ECG images in our model overcomes the implementation challenges arising from black box algorithms. The origin of risk-discriminative signals in precordial leads of ECG images suggests a left ventricular origin of the predictive signals. Moreover, the consistent observation of these predictive signals in the anteroseptal and anterior leads, regardless of the lead location on printed images, also serves as a control for the model predictions. Despite localizing the class-discriminative signals in the image to the left ventricle, heatmap analysis may not necessarily capture all the model predictive features, such as the duration of ECG segments, intervals, or ECG waveform morphologies which might have been used in model predictions. However, visual representations that are consistent with clinical knowledge could explain parts of the model prediction process and address the hesitancy in uptake of these tools in clinical practice.^24^

An important finding was the significantly increased risk of incident LV systolic dysfunction among patients with model-predicted positive screen but LVEF _≥_ 40% on concurrent echocardiography. These findings demonstrate an electrocardiographic signature that may precede the development of echocardiographic evidence of LV systolic dysfunction. This was previously reported in signal-based models,^10^ further suggesting that the detection of LV systolic dysfunction on ECG images represents a similar underlying pathophysiological process. These observations suggest a potential role for AI-based ECG models in risk stratification for future development of cardiovascular disease.^25^

Our study has certain limitations that merit consideration. First, we developed this model among patients with both ECGs and echocardiograms. Therefore, the training population selected likely had a clinical indication for echocardiography, differing from the broader real-world use of the algorithm for screening tests for LV systolic dysfunction among those without any clinical disease. The excellent performance of our algorithm across demographic subgroups and the validation population would suggest robustness and generalizability of the effects though prospective assessments in the intended screening setting are warranted. Second, the model performance may vary by degree of LV systolic dysfunction. Though we chose an LVEF threshold of 40% due to its therapeutic implications, such as an indication for disease-modifying guideline-directed medical therapies,^4^ the model identifies individuals with mild dysfunction.

This may highlight a shared signature of LV systolic dysfunction among those with LVEF<40%, and with LVEF of 40-50%, but could also represent the lack of precision of LVEF measurement by echocardiography relative to more precise approaches, such as magnetic resonance imaging.^26,27^ Third, while we incorporated four ECG formats during its development and demonstrated that the model had a consistent performance on a range of commonly used and novel layouts that were not included in the development, we cannot ascertain whether it maintains performance on every novel format. Fourth, while the model development pursues preprocessing the ECG signal for plotting images, these represent standard processes performed before ECG images are generated and/or printed by ECG machines. Therefore, any other processing of images is not required for real-world application, as demonstrated in the application of the model to the external validation sets.

## CONCLUSIONS

We developed an automated algorithm to detect LV systolic dysfunction from ECG images, demonstrating a robust performance across subgroups of patient demographics, ECG formats and calibrations, and clinical practice settings. Given the ubiquitous availability of ECG images, this approach represents a strategy for automated screening of LV systolic dysfunction, especially in resource-limited settings.

## Data Availability

All data produced in the present work are contained in the manuscript

## Author contributions

RK conceived the study and accessed the data. VS and RK developed the model. VS, AAN, LD, AK, and RK pursued the statistical analysis. VS and AAN drafted the manuscript. All authors provided feedback regarding the study design and made critical contributions to writing of the manuscript. RK supervised the study, procured funding, and is the guarantor.

## Model Availability

The model is available in an online format for research use at https://www.cards-lab.org/ecgvision-lv

## ABBREVIATIONS AND ACRONYMS

LV: Left ventricle
ECG: Electrocardiography
AI: Artificial Intelligence
LVEF: Left ventricular ejection fraction
YNHH: Yale New Haven Hospital
TTE: Transthoracic echocardiography
LRH: Lake Regional Hospital
ELSA-Brasil: ELSA-Brasil, Estudo Longitudinal de Saúde do Adulto (The Brazilian Longitudinal Study of Adult Health)
Grad-CAM: Gradient-weighted Class Activation Mapping
AUROC: Area under receiving operation characteristics
AUPRC: Area under precision recall curve

