## Supplementary material for "Detection of Left Ventricular Systolic Dysfunction from Electrocardiographic Images": Online Supplement

### **SUPPLEMENTAL MATERIAL**

**(Sangha et al.)**

**EXTERNAL VALIDATION DATA AND PROCEDURES**

The model was externally validated on ECG images obtained through three separate sampling strategies:

1- Consecutive inclusion at Cedars Sinai Medical Center in Los Angeles, CA, USA
2- Stratified convenience sampling at 4 centers, evaluating the logistics of deploying the model directly by clinicians
   a. Outpatient clinics of Yale New Haven Hospital (YNHH) across the state of Connecticut (CT), USA,
   b. Lake Regional Hospital (LRH) at Osage Beach, MO, USA,
   c. Memorial Hermann Southeast Hospital at Houston, TX, USA, and
   d. Methodist Cardiology Clinic at San Antonio, TX, USA
3- Community-based prospective sampling in Brazilian Longitudinal Study of Adult Health (ELSA-Brasil)

The sampling strategies and ECG image acquisition details are described for each center below.

**Cedars Sinai Medical Center:** ECG images were obtained during outpatient encounters of patients at Cedars Sinai Medical Center between January through March 2019. A total of 879 ECGs from unique individuals, including 99 with LVEF < 40%, were included in this set. Clinically used PDFs of ECGs were collected for model validation. These ECGs had three rhythm strips at the bottom (V1, II, and V5), which was different from the original four layouts included in the model training. This sample represents all individuals who underwent an echocardiography at Cedars Sinai Medical Center during this period. The prevalence of LV systolic dysfunction in this sample was not pre-specified, hence represents the true prevalence rate of individuals with low LVEF in this population in this interval.

**Outpatient Clinics of YNHH:** ECG images from outpatient clinics of YNHH were obtained during January through March 2022 and included 147 ECGs from unique individuals, 27 with LVEF < 40%. This was a convenience sample, with oversampling individuals with LVEF < 40% to achieve a target prevalence of 20% for LV systolic dysfunction, which was estimated to be twice as large as the underlying prevalence of LV systolic dysfunction in this population (10%). The ECG images were manually captured through image capture from electronic health record. These images had a similar layout to the standard ECG format used in model training but had the lead II rather than lead I as the rhythm strip. Moreover, there were several real-world noise artifacts in these images, including the shade of the page, vertical lines demarcating the leads, and differences in the location of the lead labels.

**Lake Regional Hospital (LRH):** LRH is a community hospital and part of a rural US hospital system in Osage Beach, MO. Data from this external set included 100 ECG images, with 43 from patients with LVEF < 40%. Individuals with LVEF < 40% were oversampled in to achieve a target prevalence of 40% for LV systolic dysfunction in this convenience sample. The ECG images in this sample had a similar layout as the standard ECG format in the train set but had lead II rather than lead I as the rhythm strip. The images were obtained through image captured from the electronic health records of individuals. There were unique noise real-world artifacts present in these images too, including a different background color, the layout of the grid over which the waveform data are displayed, as well as the location and the font of the lead label.

**Memorial Hermann Southeast Hospital:** 50 ECG images were obtained from inpatient admissions at Memorial Hermann Southeast Hospital in Houston, TX. Patients with LV systolic dysfunction were oversampled for a target prevalence of 20% in this convenience sample, which included 11 individuals with LVEF < 40% in the final sample. ECGs in this sample were in printed format and had three rhythm leads (V1, II, and V5) at the bottom The ECG paper copies in the medical records were scanned.

**Methodist Cardiology Clinic:** This dataset included ECGs from 50 individuals, including 11 individuals with LVEF < 40% from inpatient admissions or outpatient visits at Methodist Cardiology Clinic in San Antonio, TX. Individuals with LVEF < 40% were oversampled. ECGs were obtained through screenshots of electronic medical records and had several different outlines, including one (lead II), two (leads II and V1), or three (leads II, V1, or V5) leads as the rhythm strips at the bottom.

**ELSA-Brasil:** The Brazilian Longitudinal Study of Adult Health (ELSA-Brasil) studied the development and progression of clinical and subclinical chronic diseases, particularly cardiovascular diseases, and diabetes. Study participants were enrolled from the community at 5 academic centers in Brazil between 2008-2019. In this prospective study, echocardiography and ECG data were obtained from enrolled participants by protocol and not by indication. A total of 2,577 individuals, including 30 with LVEF < 40%, had ECG-echocardiography data, all of whom were included in this validation set. Notably, the prevalence of LV systolic dysfunction in this sample is lower than the other external sets (~1%). ECG images were obtained through screenshots of electronic health records.

**Table S1. Analytic packages and language used for model development and statistical analysis**

| **Programming Language/Package** | **Version** |
| --- | --- |
| Python | 3.9.5 |
| TensorFlow | 2.8.0 |
| scikit-learn | 0.24.2 |
| pandas | 1.3.1 |
| numpy | 1.19.5 |

**Table S2. Baseline characteristics of study population.** Data presented as median [IQR] for age and number (percent) for other variables.

| **Characteristic** | **Total (N = 116,210)** |
| --- | --- |
| **Sex** |  |
| Female | 59,282 (51.0%) |
| Male | 56,917 (49.0%) |
| Missing | 11 (0.0%) |
| **Age (years)** | 68 [56-78] |
| **Race** |  |
| Hispanic | 9,349 (8.0%) |
| White | 75,928 (65.3%) |
| Black | 14,000 (12.0%) |
| Other | 16,843 (14.5%) |

**Table S3. Performance of model on different image formats created from the held-out test set.** Abbreviations: PPV, positive predictive value; NPV, negative predictive value; AUROC, area under receiver operating characteristic curve; AUPRC, area under precision recall curve..

| **Format** | **PPV** | **NPV** | **Specificity** | **Sensitivity** | **AUROC** | **AUPRC** |
| --- | --- | --- | --- | --- | --- | --- |
| Standard | 0.257 | 0.988 | 0.769 | 0.892 | 0.910 (0.901 - 0.919) | 0.545 (0.513 - 0.581) |
| Two-Rhythm | 0.256 | 0.987 | 0.769 | 0.889 | 0.907 (0.897 - 0.916) | 0.533 (0.500 - 0.566) |
| Alternate | 0.246 | 0.987 | 0.756 | 0.892 | 0.908 (0.899 - 0.917) | 0.534 (0.501 - 0.567) |
| Shuffled | 0.261 | 0.987 | 0.777 | 0.882 | 0.911 (0.902 - 0.920) | 0.538 (0.504 - 0.575) |

**Table S4. Performance of model on novel image formats created from the held-out test set.** **Standard format was used both in model training and validation and is presented for comparison. The three other layouts were only used for validation to assess model performance on image formats not encountered before.** Abbreviations: PPV, positive predictive value; NPV, negative predictive value; AUROC, area under receiver operating characteristic curve; AUPRC, area under precision recall curve.

| **Format** | **PPV** | **NPV** | **Specificity** | **Sensitivity** | **AUROC** | **AUPRC** |
| --- | --- | --- | --- | --- | --- | --- |
| Standard | 0.257 | 0.988 | 0.769 | 0.892 | 0.910 (0.901 - 0.919) | 0.545 (0.513 - 0.581) |
| Three-Rhythm | 0.224 | 0.990 | 0.715 | 0.919 | 0.907 (0.898 - 0.917) | 0.533 (0.503 - 0.569) |
| No-Rhythm | 0.204 | 0.988 | 0.684 | 0.907 | 0.887 (0.877 - 0.898) | 0.465 (0.432 - 0.500) |
| Rhythm on Top | 0.220 | 0.987 | 0.715 | 0.897 | 0.901 (0.892 - 0.911) | 0.504 (0.468 - 0.540) |

**Table S5. Performance of model on standard format held-out test set images generated from differently calibrated ECGs.** Abbreviations: PPV, positive predictive value; NPV, negative predictive value; AUROC, area under receiver operating characteristic curve; AUPRC, area under precision recall curve.

| **Calibration** | **PPV** | **NPV** | **Specificity** | **Sensitivity** | **AUROC** | **AUPRC** |
| --- | --- | --- | --- | --- | --- | --- |
| 5 mm/mV | 0.179 | 0.991 | 0.615 | 0.939 | 0.898  (0.888 – 0.908) | 0.504  (0.470 – 0.538) |
| 10 mm/mV | 0.257 | 0.988 | 0.769 | 0.892 | 0.910  (0.901 – 0.919) | 0.545  (0.509 – 0.580) |
| 20 mm/mV | 0.236 | 0.981 | 0.757 | 0.838 | 0.882  (0.871 0.893) | 0.466  (0.432 – 0.500) |
| Mixed* | 0.248 | 0.988 | 0.757 | 0.895 | 0.908  (0.899 – 0.918) | 0.538  (0.503 – 0.573) |

*The mixed calibration set was generated with a partial sample of 5 mm/mV and 20 mm/mV calibrations from high and low voltages in lead I (together representing 25% of sample from test set), along with 10 mm/mV (remaining 75% of test set)

**Table S6: Performance of the model in detecting LV systolic dysfunction from ECGs stratified by PR interval.** Abbreviations: PPV, positive predictive value; NPV, negative predictive value; AUROC, area under receiver operating characteristic curve; AUPRC, area under precision-recall curve.

| **Metric** | **PR Interval ≤ 200** | **PR Interval > 200** |
| --- | --- | --- |
| **PPV** | 0.247 | 0.265 |
| **NPV** | 0.99 | 0.981 |
| **Specificity** | 0.809 | 0.73 |
| **Sensitivity** | 0.881 | 0.871 |
| **AUROC** | 0.920 (0.909 – 0.931) | 0.895 (0.871 – 0.929) |
| **AUPRC** | 0.537 (0.502 – 0.589) | 0.570 (0.495 – 0.673) |

**Table S7: Performance of the model in detecting LV systolic dysfunction from paced and non-paced ECGs.** Abbreviations: PPV, positive predictive value; NPV, negative predictive value; AUROC, area under receiver operating characteristic curve; AUPRC, area under precision recall curve.

| **Metric** | **Non-paced ECGs** | **Paced ECGs** |
| --- | --- | --- |
| **PPV** | 0.239 | 0.363 |
| **NPV** | 0.987 | 0.969 |
| **Specificity** | 0.785 | 0.317 |
| **Sensitivity** | 0.872 | 0.975 |
| **AUROC** | 0.908 (0.898 – 0.919) | 0.817 (0.784 – 0.858) |
| **AUPRC** | 0.519 (0.491 – 0.562) | 0.617 (0.554 – 0.714) |

**Table S8: Performance of model on ECGs after exclusion of atrial fibrillation, atrial flutter, conduction disorders, and paced rhythms.** Abbreviations: PPV, positive predictive value; NPV, negative predictive value; AUROC, area under receiver operating characteristic curve; AUPRC, area under precision recall curve.

| **Metric** | **Performance** |
| --- | --- |
| **PPV** | 0.218 |
| **NPV** | 0.990 |
| **Specificity** | 0.840 |
| **Sensitivity** | 0.840 |
| **AUROC** | 0.919 (0.905 – 0.933) |
| **AUPRC** | 0.536 (0.481 – 0.585) |

**Table S9: Performance of the model, stratified by patients with TTE performed before, after, or on the same day as the ECG.**

| **Performance Metric** | **TTE and ECG on the Same Day**  **N = 2276 (19.6%)** | **TTE After ECG**  **N = 6881 (59.2%)** | **TTE Before ECG**  **N = 2464 (21.2%)** |
| --- | --- | --- | --- |
| **PPV** | 0.277 | 0.232 | 0.291 |
| **NPV** | 0.983 | 0.990 | 0.985 |
| **Specificity** | 0.744 | 0.787 | 0.737 |
| **Sensitivity** | 0.882 | 0.890 | 0.905 |
| **AUROC** | 0.900 (0.881 - 0.920) | 0.920 (0.908 - 0.932) | 0.894 (0.874 - 0.915) |
| **AUPRC** | 0.536 (0.468 - 0.608) | 0.534 (0.483 - 0.582) | 0.590 (0.531 - 0.649) |

**Table S10: Confusion Matrices for Model Performance on real-world external validation datasets.** Abbreviations: EF, Ejection fraction; ELSA-Brasil, Estudo Longitudinal de Saúde do Adulto (The Brazilian Longitudinal Study of Adult Health); LRH, Lake Regional Hospital; YNHH, Yale New Haven Hospital

| **Validation Site** | **Ejection Fraction** | **Prediction Score** | | | |
| --- | --- | --- | --- | --- | --- |
|  |  | **0-0.1** | **0.1-0.2** | **0.2-0.5** | **0.5-1** |
| **Cedars Sinai Medical Center** | < 40% | 13  (13.1%) | 11  (11.1%) | 35  (35.4%) | 40  (40.4%) |
|  | 40 – 50% | 36  (37.1%) | 16  (16.5%) | 29  (29.9%) | 16  (16.5%) |
|  | > 50% | 566  (82.9%) | 54  (7.9%) | 44  (6.4%) | 19  (2.8%) |
| **Outpatient YNHH Clinics** | < 40% | 0  (0%) | 0  (0%) | 7  (25.9%) | 20  (74.1%) |
|  | 40 – 50% | 2  (14.3%) | 2  (14.3%) | 6  (42.9%) | 4  (28.6%) |
|  | > 50% | 65  (61.3%) | 13  (12.3%) | 18  (17.0%) | 10  (9.4%) |
| **LRH** | < 40% | 1  (2.3%) | 2  (4.7%) | 15  (34.9%) | 25  (58.1%) |
|  | 40 – 50% | 4  (25.0%) | 6  (37.5%) | 4  (25.0%) | 2  (12.5%) |
|  | > 50% | 17  (41.4%) | 11  (26.8%) | 12  (29.3%) | 1  (2.4%) |
| **Memorial Hermann Southeast Hospital** | < 40% | 1  (9.1%) | 0  (0%) | 1  (9.1%) | 9  (81.8%) |
|  | 40 – 50% | 5  (55.6%) | 2  (22.2%) | 1  (11.1%) | 1  (11.1%) |
|  | > 50% | 18  (60.0%) | 4  (13.3%) | 7  (23.3%) | 1  (3.3%) |
| **Methodist Cardiologist Clinic** | < 40% | 0  (0%) | 2  (18.2%) | 3  (27.3%) | 6  (54.5%) |
|  | 40 – 50% | 5  (83.3%) | 0  (0%) | 0  (0%) | 1  (16.7%) |
|  | > 50% | 21  (63.6%) | 2  (6.1%) | 8  (24.2%) | 2  (6.1%) |
| **ELSA-Brasil** | < 40% | 9  (30.0%) | 4  (13.3%) | 5  (16.7%) | 12  (40.0%) |
|  | 40 – 50% | 32  (76.2%) | 2  (4.8%) | 5  (11.9%) | 3  (7.1%) |
|  | > 50% | 2454  (98.0%) | 29  (1.2%) | 19  (0.7%) | 3  (0.1%) |

#### **Figure S1. Flow chart of study cohort and analysis.**

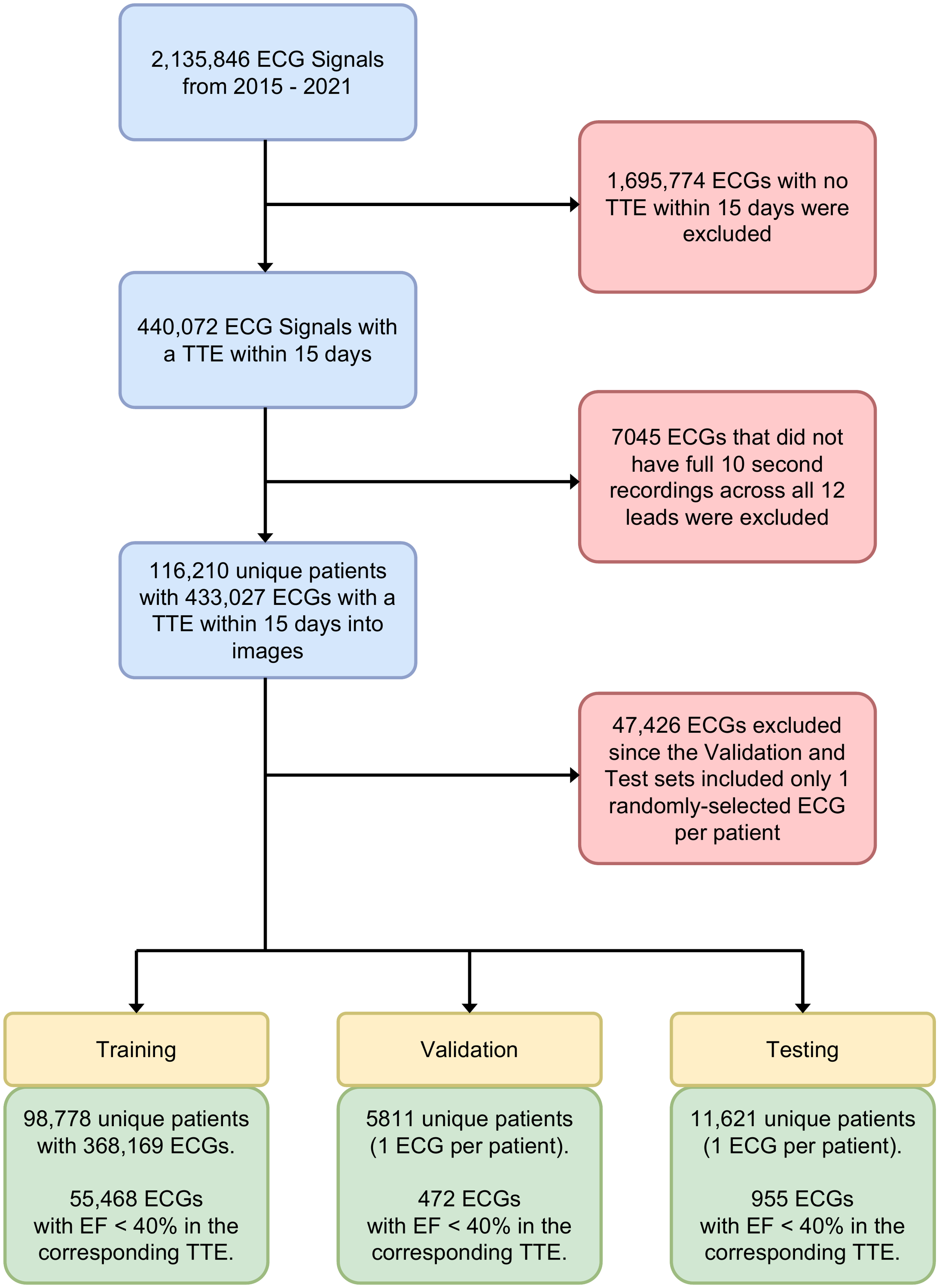

#### **Figure S2. EfficientNet-B3 architecture used for model development.** Our image-based convolutional neural network (CNN) is designed Efficientnet-B3 architecture to recognize visual patterns of LV systolic dysfunction from ECG-images. The input layer receives the ECG image as a matrix of pixel values. The convolutional layer applies a set of learnable filters to the input image that slide across the image, convolving it and producing an output feature map. These filters detect different features, such as edges, shapes, or textures, that are important for identifying patterns in the ECG image. The pooling layer reduces the spatial size of the output feature maps by performing a down-sampling operation, which helps to make the model more robust to variations in the image. Finally, the output layer produces the prediction of LV systolic dysfunction from the image.

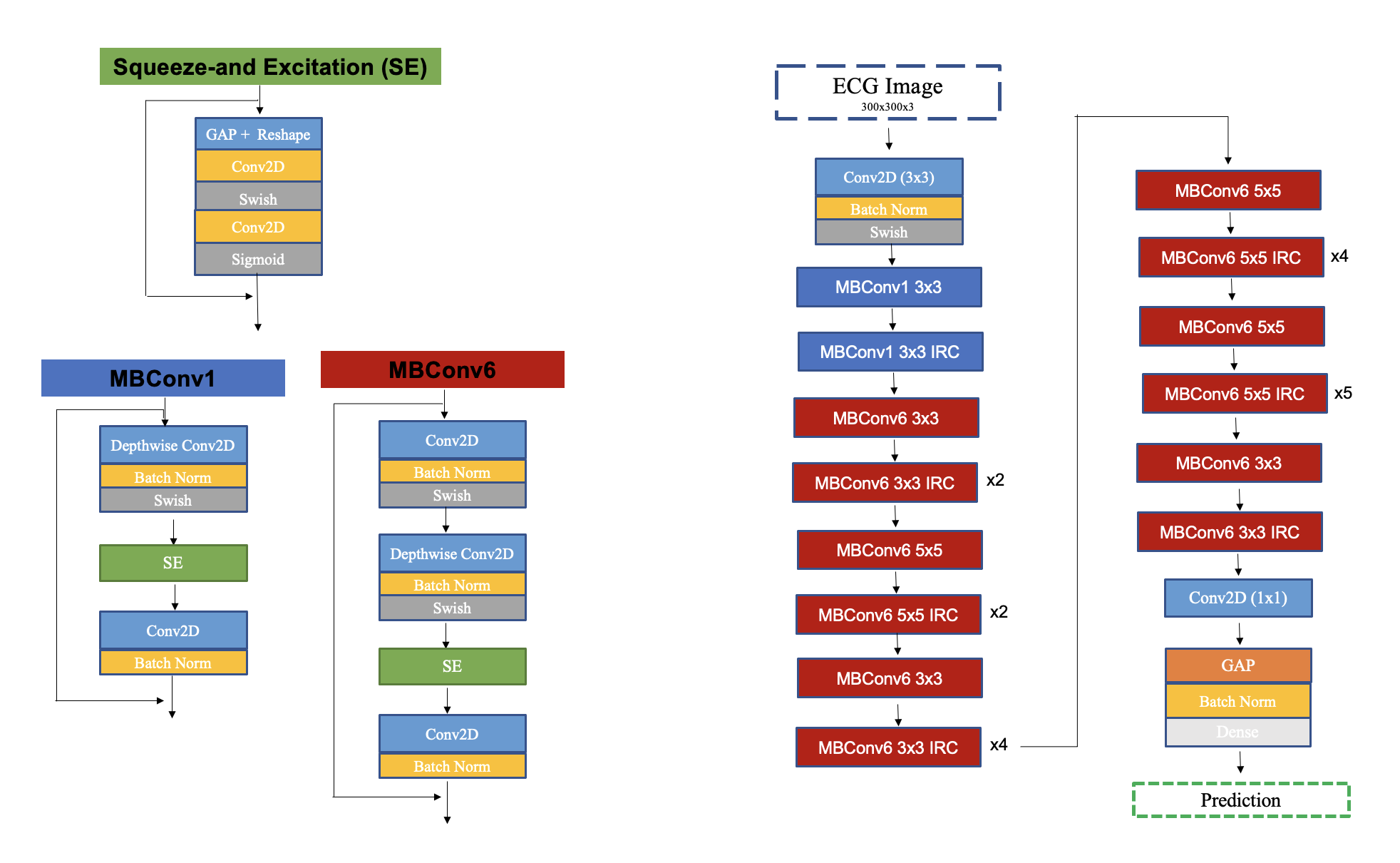

**Figure S3. Novel ECG image formats. A) standard format with lead I as rhythm strip, B) three-rhythm format with leads I, II, and V1 as rhythm strips, C) no-rhythm format with no rhythm strip, and D) rhythm on top with lead I as rhythm strip located on top. Standard format was used both in model training and validation and is presented for comparison. The three other layouts were only used for validation to assess model performance on image formats not encountered before.**

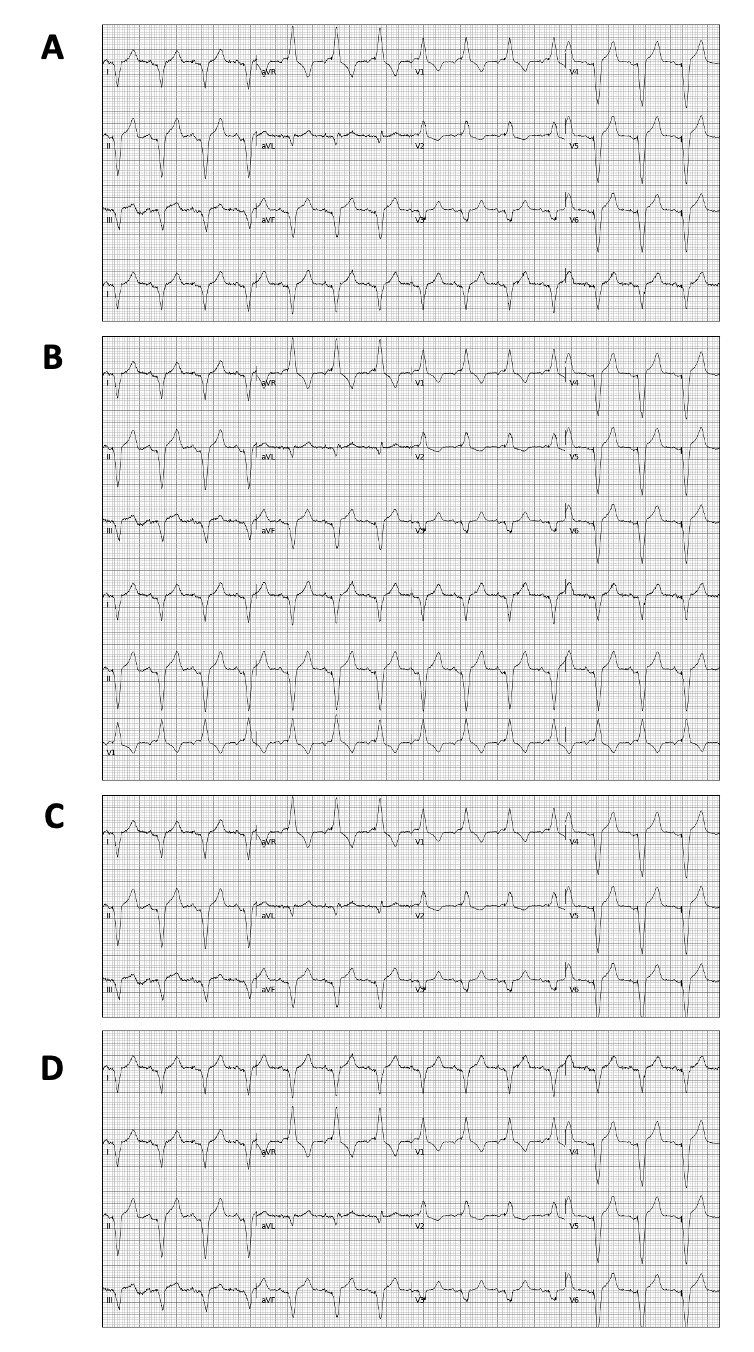

**Figure S4. ECG calibrations in validation studies. Model performance was assessed across various ECG calibrations at A) 10 mm/mV, B) 5 mm/mV, C) 20 mm/mV.**

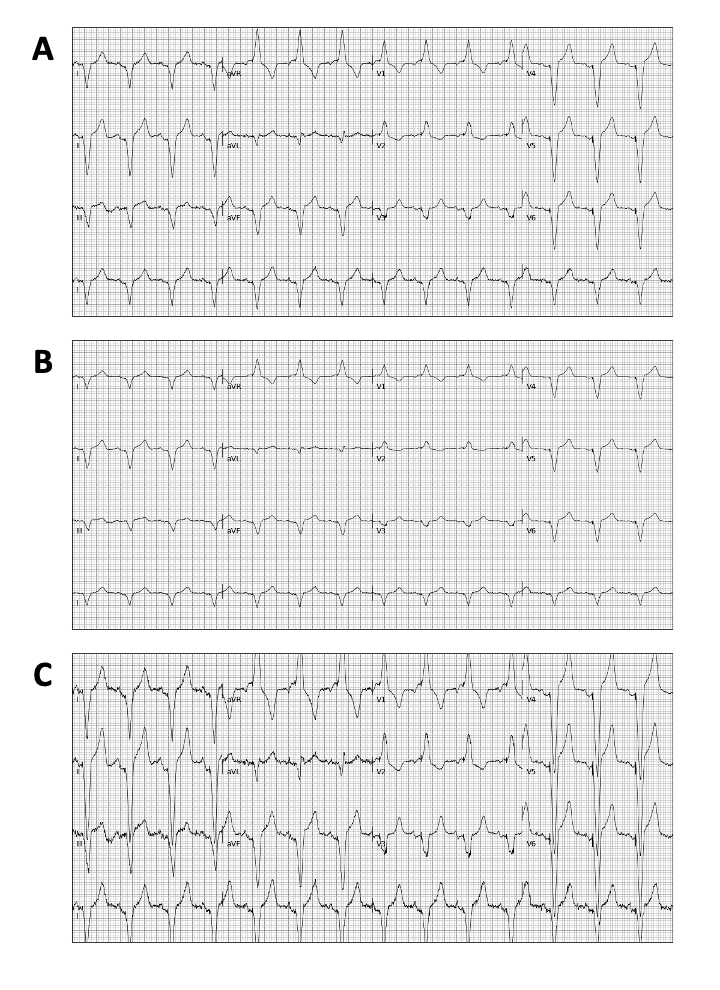

**Figure S5. Representative image of a real-world electrocardiogram from Cedars Sinai Medical Center, Los Angeles, CA used for validation.**

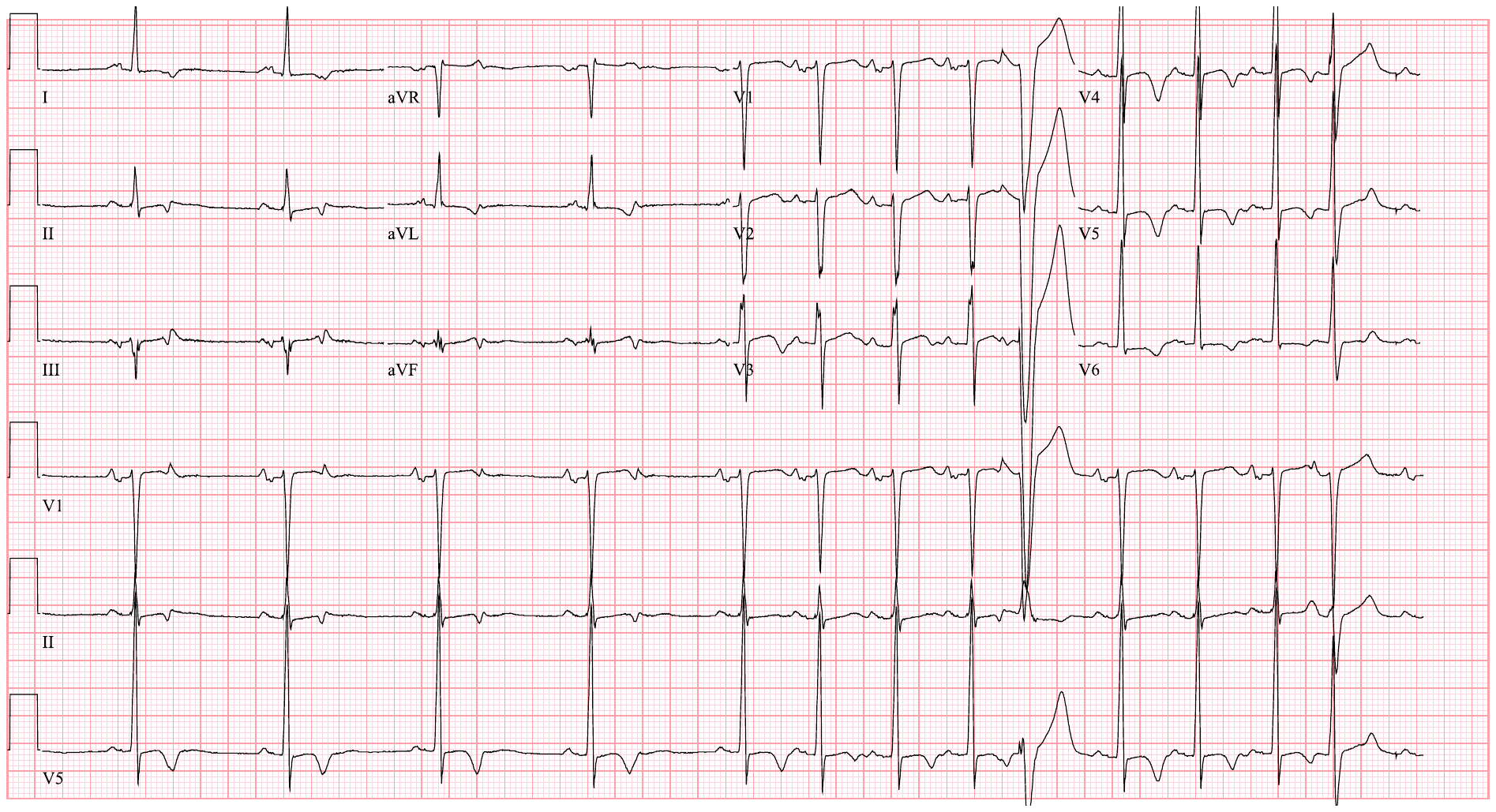

**Figure S6. Representative examples of real-world electrocardiograms from A) outpatient clinics of Yale New Haven Hospital (YNHH), and B) Lake Regional Hospital (LRH) used for validation.**

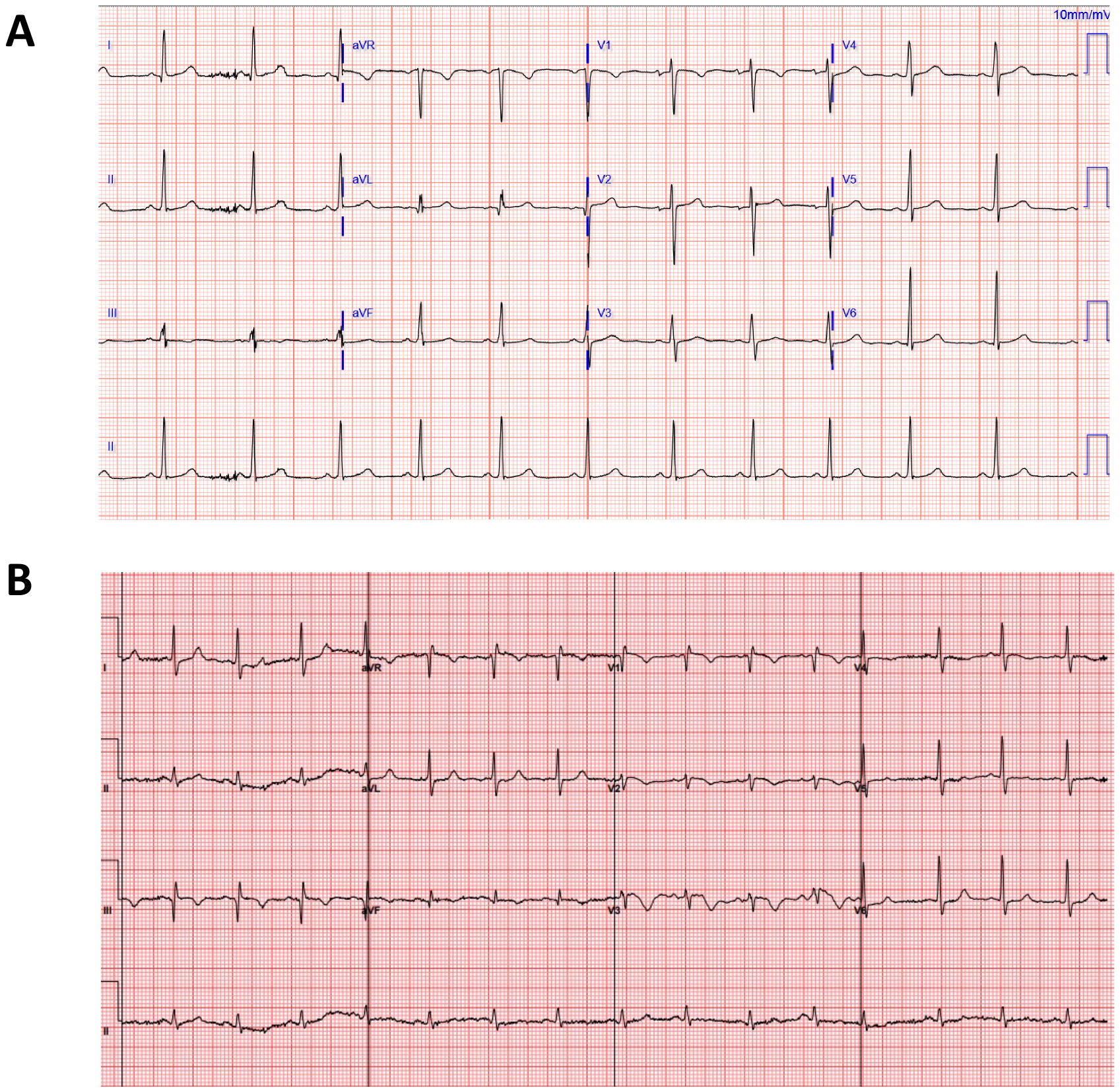

**Figure S7. Representative examples of real-world electrocardiograms from Memorial Hermann Health System in Houston, TX used for external validation.**

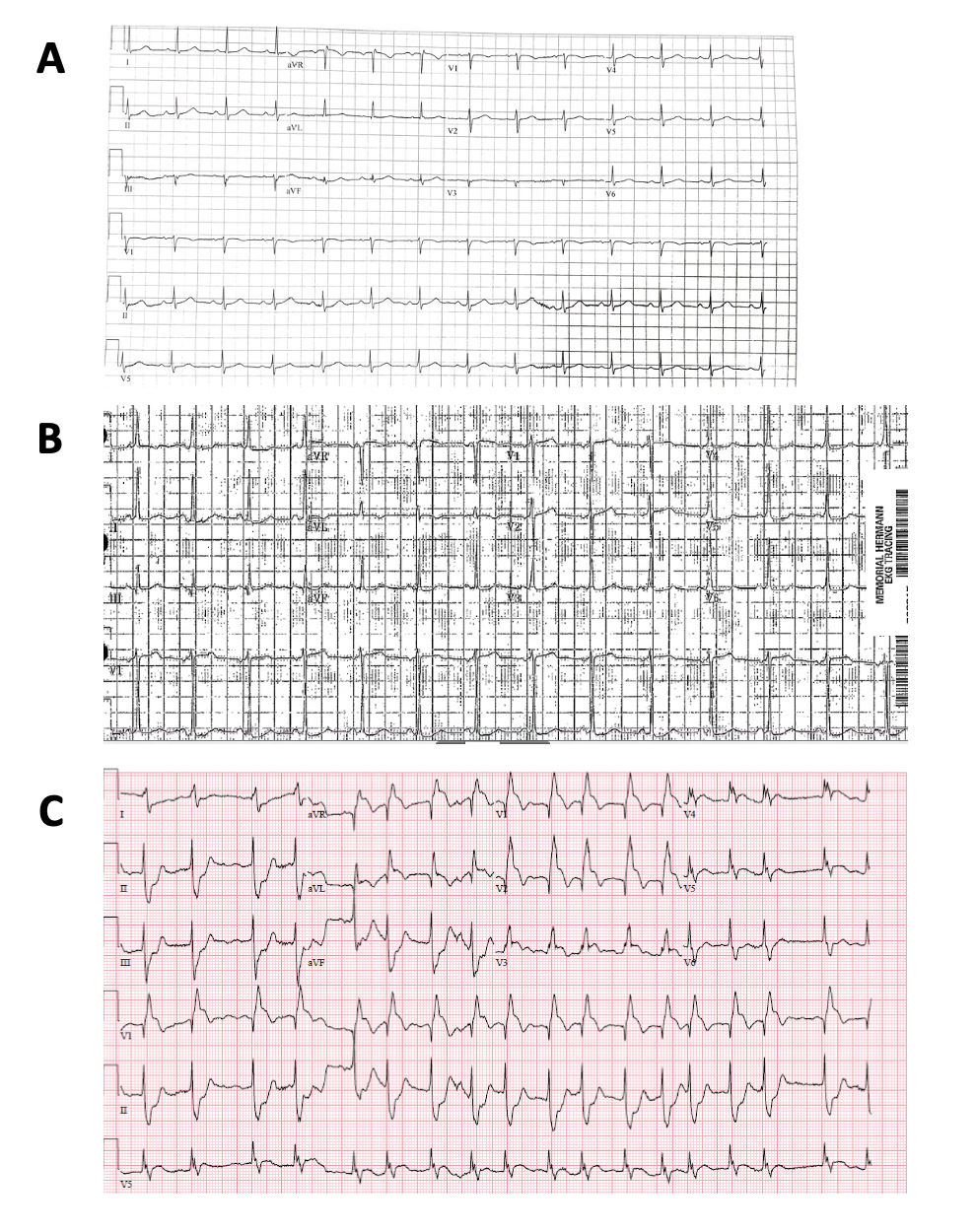

**Figure S8. Representative examples of real-world electrocardiograms from Methodist Cardiology Clinic in San Antonio, TX for external validation.**

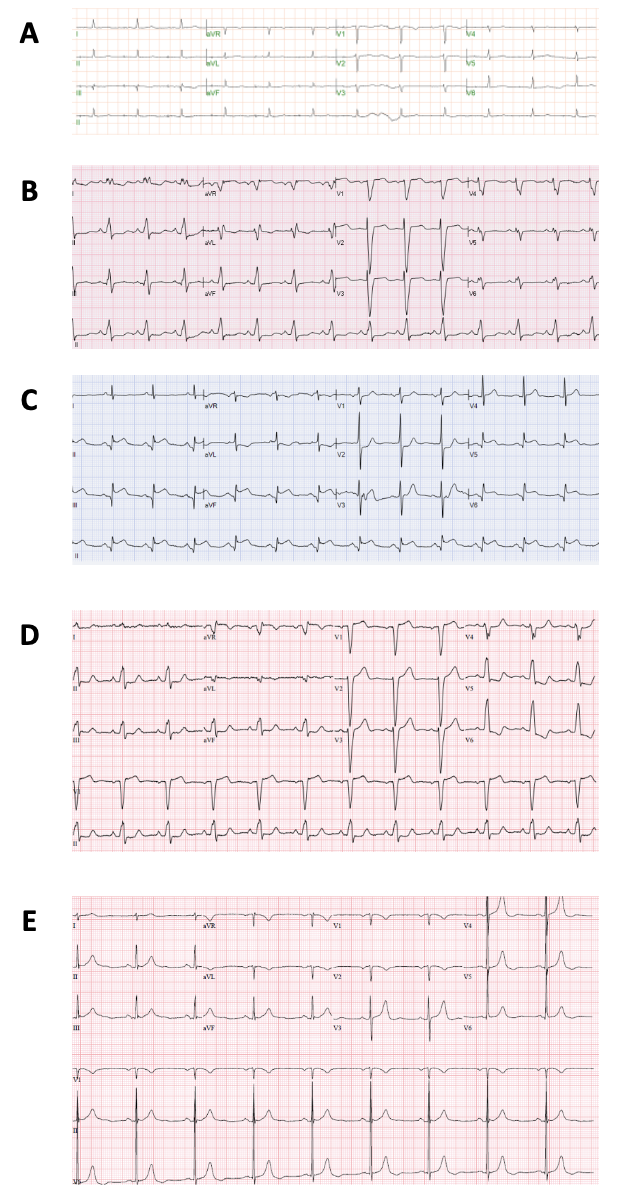

**Figure S9: (A) Proportion of individuals with LV Systolic Dysfunction across deciles of model-predicted probabilities of LV Systolic Dysfunction (B) Mean LV ejection fraction across deciles of predicted probability of LV Systolic Dysfunction.**

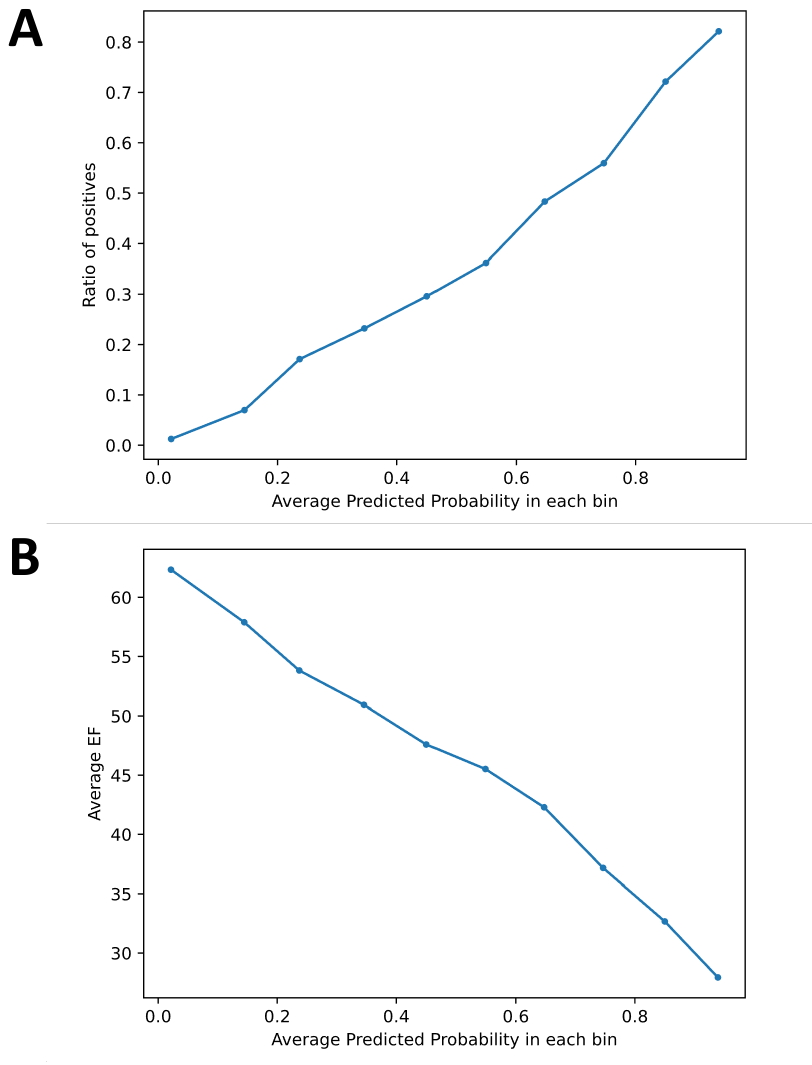

**Figure S10. Examples of Gradient-weighted Class Activation Mapping (Grad-CAM) analysis of electrocardiograms from four individuals with positive (A and B) and negative (C and D) model predictions for left ventricular systolic dysfunction. Class-discriminating signals localize to anterior leads in positive cases (A and B).**

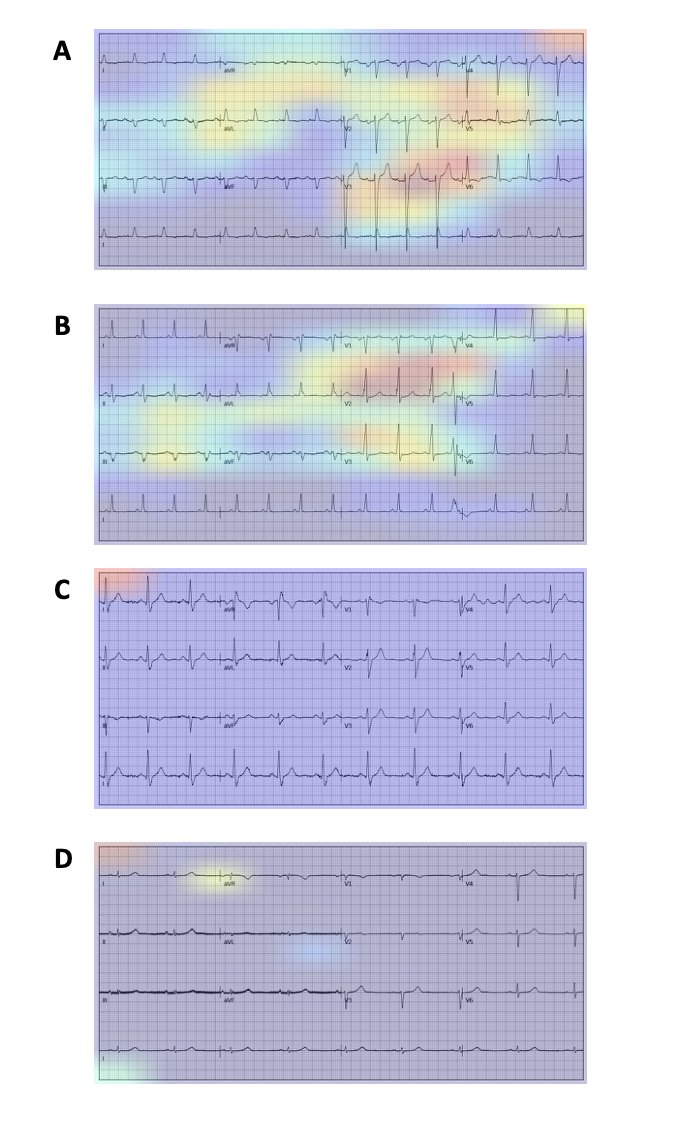

**Figure S11. Representative examples of Gradient-weighted Class Activation Mapping (Grad-CAM) analysis of electrocardiograms from each validation center A) outpatient clinics of Yale New Haven Hospital (YNHH), B) Lake Regional Hospital (LRH), C) Memorial Hermann Health, and D) Methodist Cardiology Clinic.**

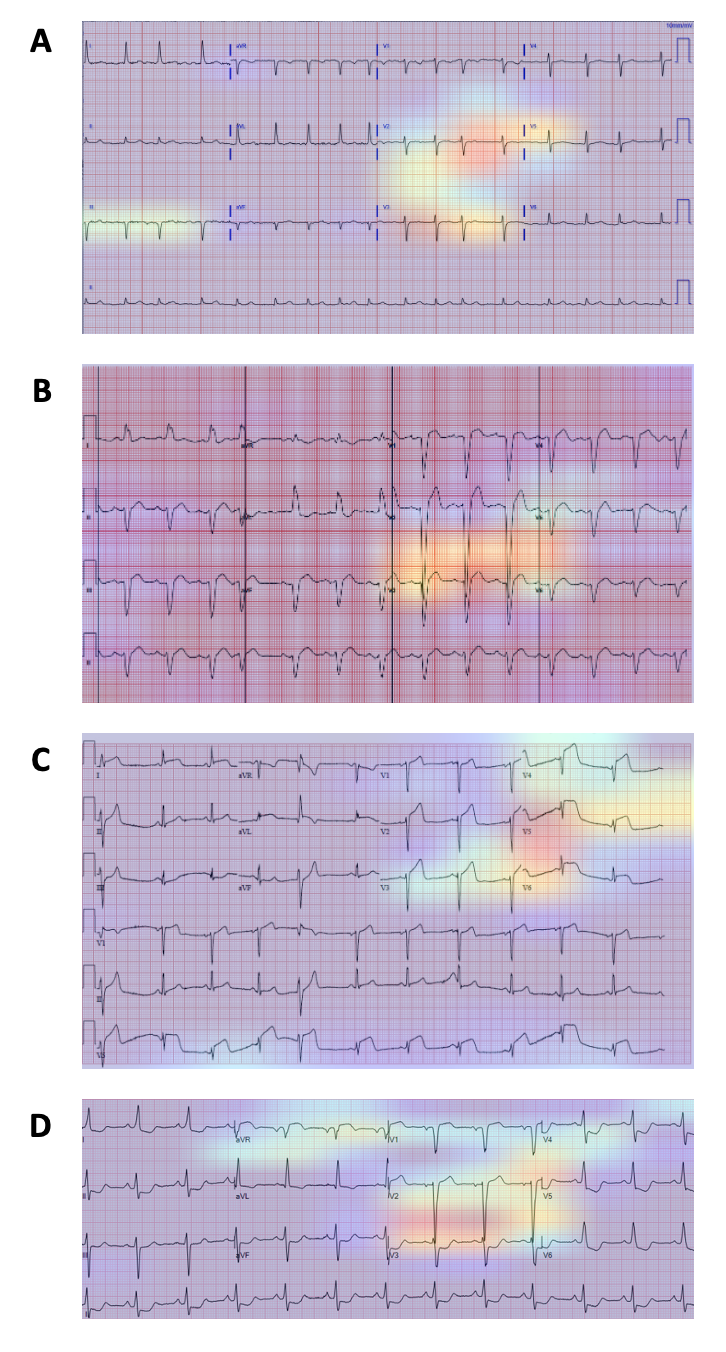
